# Reporting of patient involvement: A mixed-methods analysis of current practice in health research publications

**DOI:** 10.1101/2022.04.20.22274033

**Authors:** Sarah Weschke, Delwen Louise Franzen, Anna Karolina Sierawska, Lea-Sophie Bonde, Daniel Strech, Susanne Gabriele Schorr

**Author notes:** These authors contributed equally to this work (Sarah Weschke, Delwen Louise Franzen). **CORRESPONDING AUTHOR** Sarah Weschke; Berlin Institute of Health at Charité – Universitätsmedizin Berlin, QUEST Center for Responsible Research, Charitéplatz 1, 10117 Berlin, Germany.

## Abstract

**Objectives:** To evaluate the extent and quality of patient involvement reporting in examples of current practice in health research.

**Design:** Mixed-methods study. We used a targeted search strategy across three cohorts to identify health research publications that reported patient involvement: publications published in *The BMJ*, publications listed in the PCORI database, and publications citing the GRIPP2 reporting checklist for patient involvement or a critical appraisal guideline for user involvement. Publications were coded according to three coding schemes: “Phase of involvement”, the GRIPP2-SF reporting checklist, and the critical appraisal guideline.

**Outcome measures:** The phase of the study in which patients were actively involved. For the BMJ sample, the proportion of publications that reported patient involvement. The quality of reporting based on the GRIPP2 short form reporting guideline. The quality of patient involvement based on the critical appraisal guideline. Quantitative and qualitative results are reported.

**Results:** We included 87 publications that reported patient involvement. Patients were most frequently involved in study design (90% of publications, n=78), followed by study conduct (70%, n=61), and dissemination (40%, n=35). Reporting of patient involvement was often incomplete, e.g., only 39% of publications (n=34) reported the aim of patient involvement. While the methods (56%, n=49) and results (59%, n=51) of involvement were reported more frequently, qualitative analyses showed that reporting was often unspecific and the influence of patients’ input remained vague. Therefore, a systematic assessment of the quality and impact of patient involvement according to the critical appraisal guideline was not feasible across samples.

**Conclusions:** As patient involvement is increasingly seen as an integral part of the research process and requested by funding bodies, it is essential that researchers receive specific guidance on how to report patient involvement activities. Complete reporting builds the foundation for assessing the quality of patient involvement and its impact on research.

**PROTOCOL:** The protocol was published on the Open Science Framework: https://osf.io/vntgu/

**STRENGTHS AND LIMITATIONS:**

- A targeted search strategy was used to identify examples of patient involvement reporting in a variety of publication types and study designs in health research
- A mixed-methods approach allowed for an analysis of both the completeness and quality of patient involvement reporting
- In this study, we coded statements reporting on patient involvement in 87 health research publications that may be adapted for further use
- Reporting of patient involvement was insufficiently detailed to allow for a systematic assessment of the quality of patient involvement

## INTRODUCTION

Patients’ viewpoints should be included in clinical research as they are the most affected by it.[1] Different approaches can be used to make the outcomes of clinical research more relevant to patients. One option is to actively involve patients or patient representatives in study design, study conduct, and dissemination. Different terms are used to describe this active involvement, e.g., “Patient and Public Involvement” (PPI) or “Patient Engagement”. Patient involvement in health research varies widely and can be categorized for example according to the level or continuity of involvement, involvement in different phases of the research, or the methods applied for involvement.[1,2] Standards and principles for patient involvement focus mainly on the management of the relationship between patients and researchers.[3] These principles are important for a good collaboration, but only reflect one quality aspect of patient involvement or its impact on outcomes of clinical research. Given the efforts from patients, there is also an ethical imperative to reflect about their input in the publication, and the results and the impact of patient involvement should be evaluated and published.

Quality in clinical research is assessed with critical appraisal tools, such as the widely used risk of bias tool for randomized controlled trials (RCTs).[4,5] A critical appraisal tool to assess the quality of patient involvement was developed in 2010.[6] High quality reporting is needed to allow for critical appraisal and quality assessment. A reporting guideline GRIPP (Guidance for Reporting Involvement of Patients and Public) was developed in 2011[7] and updated in 2017 to GRIPP2.[8,9] GRIPP2 comes in two different formats: a long form (LF) for studies with patient involvement as primary focus and a short form (SF) for studies with patient involvement as secondary or tertiary focus, such as for example clinical studies being informed by an active involvement of patients.

In 2014, the British Medical Journal (BMJ) group endorsed a policy which made it a requirement to report on patient and public involvement in BMJ journals and recommends GRIPP2 as a reporting standard.[10] Price et al.[11] compared reporting of patient and public involvement (PPI) before and after the introduction of the BMJ policy. They found that while 86% of research articles included a PPI statement about one year after the introduction of the policy, only 11% actually reported PPI activities. Funding organizations are also likely to play an important role in improving the quality and reporting of patient involvement, especially as they increasingly require patient involvement in clinical research.[12]

The objective of this study was to analyse the extent and quality of patient involvement reporting and the quality of patient involvement in examples of current practice in health research. We are aware of several studies that investigated the rate and quality of reporting[13] or critically appraised patient involvement in specific domains of clinical research.[12,14–16] Overall, these previous assessments identified very few publications that reported patient involvement and/or engagement and reporting quality was sub-optimal (e.g., 0.4% of the sample described the active involvement of patients in orthopaedic research[16]). Jones et al.[15] also included studies which had patient involvement as primary focus, such as prioritizing research topics. Our scope was different: We focused on studies that actively involved patients to inform the study methodology (including dissemination) but did not have patient involvement as primary focus. We did not limit our analysis to a specific research area or experimental design (e.g., RCTs), but included three purposively selected cohorts of publications in which we expected reporting of patient involvement. We considered patients as people affected by the disease or topic, their family members, or representatives of those affected.

## METHODS

A protocol detailing the methods of this study was pre-registered in the Open Science Framework (April 28, 2020) and is openly available.[17, see also Supplement 1]

### Samples

In order to identify publications that report on patient involvement, we used a targeted search strategy in the following three samples:

1. Publications in the journal *The BMJ*, which requires reporting on patient involvement in research articles. We performed a Web of Science search (Web of Science Core Collection, March 4, 2020) to obtain all publications published in 2019 in *The BMJ* (document types: ‘Article’ or ‘Review’).
2. Publications listed in the PCORI database.[18] PCORI is a US-based organization funding patient-centred research, which continuously screens Medline via PubMed, relevant journals, and PCORI staff recommendations for publications on patient engagement in health research. We filtered for topic: example of engagement in health research; stakeholder involvement: patients; year: 2019.
3. Publications citing one of the two GRIPP2 publications[8,9] or the critical appraisal publication[6] in Dimensions.[19]

Details of the search strategies can be found in Supplement 2. All included publications across these samples were checked for additional links or references, which described patient involvement in more detail. If additional relevant documents were found (e.g., supplemental materials, previously published protocols), they were included in the sample.

### In- and exclusion criteria

The following inclusion and exclusion criteria were applied across all samples:

#### Study type

- Quantitative studies (randomized controlled trials, observational studies, etc.) were included; qualitative research studies were excluded.
- Systematic reviews and scoping reviews were included; narrative reviews were excluded.
- Mixed-method studies and those which used qualitative and quantitative methods were included if the methods were mainly quantitative.
- Protocols were excluded. If an original publication in our sample cited a protocol which provided more detailed information on patient involvement, this protocol was included as an additional document.
- Studies in which a tool was developed and tested were included. Studies in which tools/interventions/outcomes were developed but not applied were excluded.
- Comments, editorials, guidelines, consensus papers, and other publications, which did not aim to answer a research question, were excluded.

#### Patient involvement

- Publications were included if a patient involvement activity was described in at least one phase of the study, i.e., patients or patient representatives were actively involved in designing or running the study, were engaged as co-researchers, supported the dissemination of results, or had an advisory function; participating in a study as a “subject” or “participant” was not considered as sufficient to qualify as patient involvement.
- Publications were included if patient involvement was used to inform the study, but it was not the primary focus of the study. The authors’ decision whether to complete the short or long form of the GRIPP2 reporting checklist was used as an indicator of the focus of the study (if applicable).
- Publications were included if patient involvement activities had already been conducted (not only planned). The only exception was for patient involvement in dissemination activities. Given that dissemination activities often take place after a study is published, studies with planned patient involvement in dissemination activities were included.

All identified studies were screened by at least two members of the team. Discrepancies were discussed in the team until consensus was reached. Inclusion and exclusion criteria were refined during the process to accommodate for the wide variety of studies included in the sample. Deviating from the protocol, we decided to exclude all qualitative studies and mixed-methods studies with mainly qualitative methods. In most of these publications, it was difficult to distinguish between the active involvement of patients in the study and their involvement as subjects of the qualitative research.

### Coding

We coded all publications and additional documents using three coding schemes (Supplement 3):

1. Phase of involvement: Included publications had to report patient involvement in at least one of the three study phases: study design (sub-codes “research question” and “outcome measures”), study conduct, or dissemination (sub-code “co-authoring the manuscript”).
2. GRIPP2-SF[8] to assess the reporting of patient involvement.
3. Critical appraisal tool[6] to assess the quality of patient involvement.

We used an inclusive and pragmatic approach in the coding. For example, we accepted a statement such as “patients were included to inform the study design” as sufficient to describe the aim of patient involvement according to GRIPP2. A mere description of tasks was considered as sufficient to code “have the researchers discussed the nature of tasks” according to the critical appraisal tool. We also coded acknowledgement and contribution statements if these mentioned phases or activities of patient involvement. In view of assessing the quality and impact of patient involvement based on included publications, we coded statements that addressed criteria in the critical appraisal tool. However, the varying amount of detail reported across publications and samples did not allow for a systematic appraisal of the quality of patient involvement.

Two raters coded all included publications according to the coding schemes. Discrepancies were discussed until consensus was reached. If this was not possible, a third person assessed the respective passage and the team decided by majority vote.

### Analysis

Coded segments were exported from MAXQDA[20] and analysed further in Microsoft Excel. Codes from additional documents were merged with that of the original study. We quantified how many publications in each sample reported one of the codes at least once. For the BMJ sample we additionally report the frequency of patient involvement across all publications, given the journal’s requirement to report whether patient and public involvement has taken place.[10]

Additionally, we conducted a qualitative content analysis based on the extracted GRIPP2 codes. Similar codes within the same GRIPP2 category were grouped into overarching themes. All coded segments were also reviewed for illustrative examples.

### Patient and Public Involvement

Patients or the public were not involved in the planning or conduct of this meta-research study. The analyses were not restricted to studies on specific diseases or patient populations; therefore, it would not have been adequate to include a specific patient group since this research is not specifically relevant for them. The main target audience includes researchers and other stakeholders in health research (e.g., journal editors, funders). The results have been discussed in workshops with health researchers and patients and/or patient representatives and other stakeholder (e.g., funders) to raise awareness of this topic and to describe the progress of integrating patient involvement in health research.

## RESULTS

### In- and Exclusion

A total of 87 research publications were included in the analysis after applying our inclusion and exclusion criteria (see Supplement 2). From the BMJ sample, 32 of 155 research articles (21%) were included because they reported PPI activities and qualified as quantitative study. We included a further 42 publications from the PCORI sample and 13 from the Citation sample (12 citing GRIPP2 and one citing the critical appraisal tool). Most frequently applied exclusion criteria were “no patient involvement” and “no research publication”. We included 35 additional documents, which provided further information on patient involvement described in the publications.[21]

### Phase of Involvement

Patients were most frequently involved in the study design (90% of included publications, n=78), followed by study conduct (70%, n=61), and dissemination (40%, n=35) (Table 1;[21]). In 17% (n=15) of the publications, patients were involved in formulating the research question and in 31% (n=27) in defining outcome measures.

**Table 1:** Phase of Involvement.

| Phase of Involvement | BMJ (N=32) | PCORI (N=42) | Citation (N=13) | Total (N=87) |
| --- | --- | --- | --- | --- |
| **Study design** | 75% (n=24) | 100% (n=42) | 92% (n=12) | 90% (n=78) |
| **Research question** | 16% (n=5) | 14% (n=6) | 31% (n=4) | 17% (n=15) |
| **Outcome measures** | 22% (n=7) | 40% (n=17) | 23% (n=3) | 31% (n=27) |
| **Study conduct** | 56% (n=18) | 74% (n=31) | 92% (n=12) | 70% (n=61) |
| **Dissemination** | 41% (n=13) | 36% (n=15) | 54% (n=7) | 40% (n=35) |
| **Co-authoring the manuscript** | 6% (n=2) | 21% (n=9) | 38% (n=5) | 18% (n=16) |

### GRIPP2 (short form)

Between 13% (n=4, BMJ sample) and 77% (n=10, Citation sample) of the publications reported the aim of patient involvement (Table 2;[21]). The predominant code for aim identified in our sample was ensuring that patients’ perspectives were taken into account. Coded segments ranged from vague statements (Table 2, Example (E) 1) to more elaborated accounts (Table 2, E2). Other examples included support with recruitment or the dissemination of the results and ensuring the accessibility or acceptability of the study.

**Table 2:** Reporting of patient involvement according to GRIPP2-SF: quantitative and qualitative results.

| GRIPP2 | BMJ<br>(N=32) | PCORI<br>(N=42) | Citation<br>(N=13) | Total<br>(N=87) | Examples (E) from the publications |
| --- | --- | --- | --- | --- | --- |
| **Aim**<br>Report the aim of PPI in the study | 13%<br>(n=4) | 48%<br>(n=20) | 77%<br>(n=10) | 39%<br>(n=34) | **E1: vague, unspecific statement**<br>"He was recruited to provide a patient's perspective" [23]<br>**E2: elaborated, informative statement**<br>"To ensure an improved completion rate in the full study, we hired a patient partner (B.G.S.) to approach bedside nurses, enrol participants and assist family caregivers with the completion of the questionnaires. We expect this will increase the completion rate because this family caregiver has the lived experience, perseverance and communication skills that are necessary to make the consent process less overwhelming." [24] |
| **Methods**<br>Provide a clear description of the methods used for PPI in the study | 28%<br>(n=9) | 67%<br>(n=28) | 92%<br>(n=12) | 56%<br>(n=49) | **E3: broad statement lacking details**<br>"Patient discussion groups were used as a means of involving patients in setting the research question and for determining the outcome measures." and "The outcome measures were identified after discussion groups in our unit, with patients who had previously undergone knee arthroplasty surgery." [25]<br>**E4: detailed, informative statement**<br>"During the pre-award period, the researchers developed a stakeholder management plan ("roadmap") outlining where stakeholders could influence the study. (...) Creating a stakeholder roadmap allowed us to target defined actions to engage stakeholders quickly, even before the study begun. Our stakeholder engagement plan focused on 4 key sets of activities: (1) study planning (including study design, intervention design and procedures, outcomes measurement, materials); (2) hospital/patient recruitment and retention; (3) study implementation; and (4) translation, including interpreting study findings and disseminating results back to participating communities and the public." [26] |
| **Study results**<br>Report the results of PPI in the study, including both positive and negative outcomes | 28%<br>(n=9) | 76%<br>(n=32) | 77%<br>(n=10) | 59%<br>(n=51) | **E5: broad statement lacking details**<br>"Their input helped to refine the research question and to improve the protocol considerably." [27]<br>**E6: informative statement**<br>"The group stated that participant-facing material (participant information leaflet and invitation brochure) were not inclusive enough for all 'father figures' (e.g., stepfathers). The material in question was changed to be more inclusive. To boost recruitment to the trial, the PPI group suggested organisations that might be accessed by our target population and that might be interested in supporting the study (e.g., youth clubs, after-school clubs and sports centres). Therefore, we approached and successfully recruited fathers from after-school and martial arts clubs and Scouts groups for the cultural adaptation phase. When asked about dissemination of study results, PPI representatives thought that it was important to report the findings back to the local authorities that funded the HDHK programmes and to include the use of the local authorities' social media channels for dissemination. The group encouraged the research team to be fully open about the challenges faced in delivering the HDHK programme and the research study." [28] |
| **Discussion and conclusions**<br>Comment on the extent to which PPI influenced the study overall. Describe positive and negative effects. | 16%<br>(n=5) | 52%<br>(n=22) | 69%<br>(n=9) | 41%<br>(n=36) | **E7: statement reporting on influence on usability**<br>"We enhanced the usability of our findings by engaging key stakeholders (i.e. clinicians, parents and researchers) as part of our integrated knowledge translation activities (...). Their guidance ensured that the review findings were both clinically meaningful and family-centred." [29]<br>**E8: statement providing details on the influence on the primary outcome**<br>"Patient prioritization of meaningful physician discussion over reducing anxiety and depression, therefore, resulted in a substantial redesign of both the ACP video and its evaluation trial. The team negotiated with its funder to change the primary outcome of the video's evaluation from anxiety and depression to having a meaningful discussion. Meaningfulness of the discussion was operationalized by the measure 'patient-centered nature of the patient-surgeon conversation' in the resulting video evaluation trial, procedures for which have been previously described. This change in outcome also required that language be added to the video to encourage open communication between the patient and surgeon. In the practice-oriented context of the current study, using both engagement and research approaches in endpoint selection provided an innovative means of identifying and prioritizing endpoints." [30] |
| **Reflections/critical perspective**<br>Comment critically on the study, reflecting on the things that went well and those that did not, so others can learn from this experience | 3%<br>(n=1) | 19%<br>(n=8) | 77%<br>(n=10) | 22%<br>(n=19) | **E9: statement reflecting on organizational issues**<br>"Originally, we intended to conduct these sessions in a group format, due to difficulties with PPI partners' schedule commitments, one-to-one sessions were conducted." and "It is extremely important that researchers plan PPI at the grant proposal stage and estimate the costs appropriately. If these costs are not correctly estimated during the initial stages of developing research proposals, they may cause a financial burden on PPI partners." [31]<br>**E10: statement reflecting on representativeness**<br>"One limitation will be engaging patients and family members who are deeply committed to the project and who consequently may not provide a representative range of patient-family perspectives. However, their insights will be vital to identifying key aspects of patient/family-centered decision aids and they will directly inform the next stage of our research—cognitive interviews with patients and their families about advance care planning aids." [32] |

More than half of the publications (56%, n=49) provided some information about the methods used for patient involvement in the study. However, these accounts were often not very detailed. The most predominant code was consultation or giving feedback, indicated by describing the group involved: patient representative, advisory group, patient group, adviser to the steering committee, patient engagement group, or lay representative. In many cases, even very basic information such as the number of involved patients, the frequency of meetings, or explanations on how discussions took place was lacking (Table 2, E3). Some publications reported on approaches they used for the consultation, such as working on an online platform or group meetings. Others used additional methods such as focus and discussion groups or interviews with patients to get further input on specific questions. A detailed, informative example was the development of a “roadmap” prior to the study (Table 2, E4), which served to identify how stakeholders could influence the study.

Of all publications, 59% (n=51) reported the results of patient involvement in the study. Examples of the reported outcomes included making materials easily understandable, support with or sharing ideas on recruitment, raising awareness about the study, and identifying patient-centred outcomes. The level of reporting varied from broad statements with only few or no concrete examples (Table 2, E5) to more detailed information on the outcomes of the patient involvement and its influence on the study (Table 2, E6, see also[22] for detailed information in a supplement).

Of all publications, 41% (n=36) provided information on the influence of patient involvement on the study (discussion and conclusion). Examples include influence on the intervention, recruitment, retention, usability of study findings (Table 2, E7), and outcomes (Table 2, E8).

A relatively small number of publications (22%, n=19) reported reflections and critical perspectives on patient involvement. Some reflections related to the research context and how its structure and settings may not always be welcoming for patient involvement (Table 2, E9). Others discussed a possible lack of representativeness or diversity in the sample of PPI contributors (Table 2, E10).

### Critical appraisal tool

The critical appraisal tool[6] focuses on the quality and impact of user involvement in research (Table 3;[21]). Specific appraisal criteria were rarely reported, such as discussing the level of involvement (7%, n=6), considering whether findings were disseminated appropriately to recipients (7%, n=6), or conducting a formal evaluation (6%, n=5). More general appraisal criteria were reported more frequently, such as the nature of tasks patients were asked to perform (45%, n=39), how findings were disseminated (not requiring an active part of patients) (46%, n=40), or a general evaluation of the added value of involving patients in the research process (47%, n=41).

**Table 3:** Reporting of patient involvement according to the Wright et al. (2010) critical appraisal tool: quantitative results.

| Question | Consider the following | BMJ<br>(N=32) | PCORI<br>(N=42) | Citation<br>(N=13) | Total<br>(N=87) |
| --- | --- | --- | --- | --- | --- |
| **Planning and project design** |  | **38%**<br>**(n=12)** | **52%**<br>**(n=22)** | **69%**<br>**(n=9)** | **49%**<br>**(n=43)** |
| 1. Is the rationale for involving users clearly demonstrated? | (a) Have the researchers explained the rationale for user involvement? [Rationale] | 3%<br>(n=1) | 33%<br>(n=14) | 46%<br>(n=6) | 24%<br>(n=21) |
| 2. Is the level of user involvement appropriate? | (a) Have the researchers explained and justified the level of user involvement [Level of involvement] | -- | 7%<br>(n=3) | 23%<br>(n=3) | 7%<br>(n=6) |
|  | (b) Have the researchers discussed the nature of tasks users were asked to perform (e.g., identifying the research question, selecting the research method, commenting on information sheets, data collection, data analysis, dissemination?) [Nature of tasks] | 34%<br>(n=11) | 45%<br>(n=19) | 69%<br>(n=9) | 45%<br>(n=39) |
| **Recruitment and training** |  | **6%**<br>**(n=2)** | **38%**<br>**(n=16)** | **77%**<br>**(n=10)** | **32%**<br>**(n=28)** |
| 3. Is the recruitment strategy appropriate? | (a) Have the researchers explained how users have been identified? [Identification] | 6%<br>(n=2) | 29%<br>(n=12) | 62%<br>(n=8) | 25%<br>(n=22) |
|  | (b) Have attempts been made to involve a wide cross-section of interests where appropriate (e.g., ethnic minorities, age, gender)? [Diversity] | -- | 10%<br>(n=4) | 8%<br>(n=1) | 6%<br>(n=5) |
|  | (c) Have the researchers discussed the credentials of the users involved? (E.g., Do the researchers discuss why the users involved are appropriate to meeting the aims of the involvement activity?) [Credentials] | 3%<br>(n=1) | 14%<br>(n=6) | 23%<br>(n=3) | 11%<br>(n=10) |
| 4. Is the nature of training appropriate? | (a) Have the researchers discussed the nature of the training provided? [Nature of training] | -- | 5%<br>(n=2) | 38%<br>(n=5) | 8%<br>(n=7) |
|  | (b) Is the nature and extent of the training justified by the researchers? (e.g., Do the researchers discuss how the training meets the needs of the users during the course of the study?) [Justification of the training] | -- | -- | 31%<br>(n=4) | 5%<br>(n=4) |
|  | (c) Has an account been given of user involvement training for professional researchers, where necessary? [User involvement training for researchers] | -- | 2%<br>(n=1) | -- | 1%<br>(n=1) |
| **Data collection and analysis** |  | **3%**<br>**(n=1)** | **10%**<br>**(n=4)** | **31%**<br>**(n=4)** | **10%**<br>**(n=9)** |
| 5. Has sufficient attention been given to the ethical considerations of user involvement and how these were managed? | (a) Do the researchers discuss ethical issues relating to the involvement of users in research (e.g., fatigue, the emotional demands of data collection)? [Ethical issues] | -- | 2%<br>(n=1) | 15%<br>(n=2) | 3%<br>(n=3) |
|  | (b) Are there any discussions about the management of ethical issues (e.g., provision of adequate information about research tasks, peer supervision)? [Management of ethical issues] | 3%<br>(n=1) | 2%<br>(n=1) | 8%<br>(n=1) | 3%<br>(n=3) |
| 6. Has sufficient attention been given to the methodological considerations of user involvement and how these were managed? | (a) Have the researchers discussed methodological issues relating to user involvement in research (e.g., potential impact on the quality of the data)? [Methodological issues] | -- | 10%<br>(n=4) | 8%<br>(n=1) | 6%<br>(n=5) |
|  | (b) Do the researchers discuss how methodological issues are managed (e.g., how differences in interpretations of qualitative data are negotiated)? [Management of methodological issues] | -- | 5%<br>(n=2) | 15%<br>(n=2) | 5%<br>(n=4) |
| **Dissemination** |  | **84%**<br>**(n=27)** | **43%**<br>**(n=18)** | **69%**<br>**(n=9)** | **62%**<br>**(n=54)** |
| 7. Have there been any attempts to involve users in the dissemination of findings? | (a) Have users been involved in the writing of the ...funding application? [Writing of the funding application] | 9%<br>(n=3) | 10%<br>(n=4) | 38%<br>(n=5) | 14%<br>(n=12) |
|  | .. of the publication? [Writing of the publication] | 6%<br>(n=2) | 24%<br>(n=10) | 46%<br>(n=6) | 21%<br>(n=18) |
|  | (b) Have the researchers described how the findings have been disseminated to participants and service users? [Description of dissemination] | 81%<br>(n=26) | 26%<br>(n=11) | 23%<br>(n=3) | 46%<br>(n=40) |
|  | (c) Are findings disseminated appropriately where necessary (e.g., translation of findings into different languages, provision of interim findings to participants in receipt of palliative care)? [Accessible dissemination] | 9%<br>(n=3) | 5%<br>(n=2) | 8%<br>(n=1) | 7%<br>(n=6) |
| **Evaluation and impact assessment** |  | **19%**<br>**(n=6)** | **57%**<br>**(n=24)** | **85%**<br>**(n=11)** | **47%**<br>**(n=41)** |
| 8. Has the 'added-value' of user involvement been clearly demonstrated? | (a) Do the researchers discuss what difference involving users in the design and conduct of the research has made to the research process? (I.e., Have the researchers considered whether the study and findings would look any different if users were not involved?) [Difference made to the research process] | 19%<br>(n=6) | 57%<br>(n=24) | 85%<br>(n=11) | 47%<br>(n=41) |
|  | (b) Do the researchers support the claims for the benefits of user involvement with examples from the research project? [Examples of assessment of the benefits] | 13%<br>(n=4) | 50%<br>(n=21) | 62%<br>(n=8) | 38%<br>(n=33) |
| 9. Have there been any attempts to evaluate the user involvement component of the research? | (a) Have the researchers discussed the evaluation of the impact of user involvement on the research project (e.g., impact on the length of the study, the financial cost of involvement activities, cost-benefit analyses)? [Evaluation of the made impact] | -- | 7%<br>(n=3) | 15%<br>(n=2) | 6%<br>(n=5) |
|  | (b) Do the researchers support claims about the impact of user involvement with examples from the evaluation? [Examples of evaluation] | -- | 2%<br>(n=1) | 8%<br>(n=1) | 2%<br>(n=2) |

Further appraisal criteria that were addressed in very few publications were the nature of training of patients (8%, n=7) and researchers (1%, n=1), or ethical (3%, n=3) or methodological (6%, n=5) considerations and how these were managed (3%, n=3 and 5%, n=4, respectively).

## DISCUSSION

We analysed a sample of 87 publications in health research that reported on patient involvement. While many publications provided information on general aspects relating to patient involvement, even very basic details were often lacking. For example, nearly all publications reported generally that patient involvement took place during study design. However, more specific information about whether this involvement in study design included defining the research question or prioritizing outcome measures was reported to a much lower extent. Similarly, 39% and 56% of publications reported on GRIPP2 aims and methods, respectively, but the reporting was often sub-optimal and statements rather vague. Despite authors alluding to many aspects of patient involvement included in the GRIPP2-SF and critical appraisal guidelines, we identified a need to improve completeness and details of reporting. This corroborates findings from previous studies on reporting of patient involvement and/or engagement.[12,13,16]

Due to this incomplete reporting, coding according to the GRIPP2-SF categories was a challenge: the sparseness of reporting in many of the publications meant that these categories were relatively broad and overlapping. For example, a statement such as “patients helped with the identification of meaningful outcomes” could describe the method (i.e., focusing on the process) or the results (i.e., identifying outcomes) of patient involvement. While the GRIPP2-SF reporting checklist is certainly useful to guide reporting in studies not having patient involvement as primary focus, our findings suggest that complementary measures could further bolster its impact on the quality and consistency of the patient involvement evidence base. Such measures could for example include broader requirements to include a statement on patient involvement in publications, more specific guidance for authors and peer-reviewers, and standardised formats without word count restrictions to support more complete and consistent reporting. High quality reporting is the basis for assessing the quality of patient involvement.

We observed considerable differences between our three samples regarding reporting, with both the Citation and PCORI sample providing more information on patient involvement than the BMJ sample. This is not surprising given the expected emphasis on patient involvement in the former samples compared to providing information in a mandatory section.

For the BMJ sample, we assessed the percentage of publications that reported patient involvement in the mandatory PPI section. Of all research articles published in 2019 (*n* = 155), 21% reported patient involvement activities. In the sample of Price et al.[11], which included research articles published between June 2015 and May 2016, patient involvement was reported in only 11% of the articles. Thus, the proportion of research articles reporting on patient involvement doubled in only a few years, demonstrating the impact of this journal policy to enhance visibility and to raise awareness for patient involvement. While this trend is encouraging, descriptions of patient involvement in the BMJ sample were generally very short and did not elaborate on the results of patient involvement or provide a thorough description of the process. For example, several publications reported on the inclusion of patient discussion groups without describing the composition, specific tasks, or influence of this group.

In contrast, in the PCORI and Citation samples we often found very detailed descriptions of patient involvement, including the nature of performed tasks, concrete examples of its influence on the study, and critical reflections [see 21 and table 2]. In many cases, these descriptions were provided in additional documents. This suggests that the word count limit imposed by journals likely contributes to the limited detail in patient involvement reporting. Additional documents or structured tables for reporting of patient involvement may be helpful. However, this approach may come with the risk that patient involvement is seen as an add-on rather than as an integral part of the conducted research.

### Strengths and limitations

One of the strengths of our study was the use of a targeted search strategy to identify examples of patient reporting in current practice across a variety of publication types and study designs in health research. Moreover, coding according to three distinct schemes allowed us to capture different aspects of relevance, including the phase of patient involvement, the use of and adherence to reporting guidelines (GRIPP2-SF), and the quality and impact of patient involvement (critical appraisal tool). All statements reporting patient involvement analysed in this study are openly available[21] for further use. For example, coded statements may inform the development of automated tools to detect reporting of patient involvement in publications.

We could not systematically assess the quality of patient involvement according to the critical appraisal criteria as originally planned. Quality assessment highly depends on reporting completeness and detail, which was inconsistent across publications. Such an analysis in our diverse sample might have favoured long vs. short reports, or participatory health research approaches vs. PPI activities informing a clinical trial.

Initially, we did not plan to exclude publications applying qualitative research methods. However, we did not find a clear definition to differentiate between the active involvement of stakeholders and their involvement as participants in qualitative research, for example in focus groups or interview studies.[e.g., 33–35] In some cases, both were reported in the same publication.[e.g., 33] This particular challenge has previously been noted in the context of assessing reporting of patient and public involvement.[11] Despite attempts to delineate these approaches[36], this is not common practice yet. More generally, excluded qualitative research studies that reported patient involvement often had patient involvement as primary focus. An analysis of these studies was beyond our scope.

We used GRIPP2 as a reporting guideline to assess the completeness of reporting of the included publications. The use of reporting guidelines without modification to serve as evaluation tools has been questioned by Logullo et al.[37] as their purpose is to guide writing. However, the authors of GRIPP2 explicitly stated that it can also be used for planning patient involvement or for quality assurance.[8]

## Conclusion

Despite important developments in the last years, patient involvement is still not a well-established approach in clinical or health research.[14,38] Therefore, we would encourage journals to request an obligatory patient involvement statement from their authors, and to give guidance on detailed reporting in a structured table or additional document. We would also encourage journals and funding organizations to support the reporting of patient involvement by requiring the use of GRIPP2-SF as a reporting tool. Finally, we encourage researchers to include sufficient detail on patient involvement in their study to allow others to derive and apply lessons learned in their own studies.

We expect that patient involvement will become more important in the next years to increase the relevance of research, in line with increasing demand from funders, publishers, and society. Broader implementation of policies and more specific guidance are needed to leverage the impact of existing reporting guidelines, and thereby improve the quality of the patient involvement evidence base. Complete reporting builds the foundation of assessing the quality and appropriateness of patient involvement and is essential towards increasing its impact on research.

## Data Availability

All data produced are available online at https://doi.org/10.5281/zenodo.6245751

## AUTHOR CONTRIBUTIONS

**SW:** Conceptualization, Investigation, Methodology, Project administration, Validation, Writing – original draft, Writing – review & editing; **DLF:** Conceptualization, Data curation, Investigation, Methodology, Project administration, Validation, Visualization, Writing – review & editing; **AKS:** Conceptualization, Investigation, Methodology, Validation, Visualization, Writing – review & editing; **LSB:** Investigation, Writing – review & editing; **DS:** Conceptualization, Methodology, Writing – review & editing; **SGS:** Conceptualization, Formal analysis, Investigation, Methodology, Project administration, Supervision, Validation, Visualization, Writing – review & editing.

## ACKNOWLEDGEMENT

We thank Lauren Fayish from PCORI for providing the dataset from the Engagement in Health Research Literature Explorer.

## SUPPLEMENTS

- Supplement 1: Protocol
- Supplement 2: Targeted Search and Flowchart
- Supplement 3: Coding Schemes

## DATA AVAILABILITY

The data generated in this study is openly available in Zenodo.[21; https://doi.org/10.5281/zenodo.6245751] This includes coded statements reporting on patient involvement across all publications included in this study.

