## Supplements_1-3 for "Reporting of patient involvement: A mixed-methods analysis of current practice in health research publications"

The protocol is also available at https://osf.io/vntgu/

### Protocol: Reporting and quality of patient engagement: Status quo in best practice examples

QUEST Center
Berlin Institute of Health
Charité - Universitätsmedizin Berlin
Anna-Louisa-Karsch-Str. 2
10178 Berlin
Germany

#### Corresponding Author:

Anna Sierawska:


#### Background

There is strong agreement that patients' viewpoints should be included in clinical research as they are the most affected by it (1). Different approaches can be used to make the outcomes of clinical research more relevant to patients. One option is to actively involve patients or patient representatives in study design, study conduct, and dissemination. In the present study, this is described by the term patient engagement in clinical research. Patient engagement in clinical research varies widely and can be categorized for example according to the level or continuity of engagement, engagement in different phases of the research, or the methods applied for engagement (1,2). Standards and principles for patient engagement focus mainly on the management of the relationship between patients and researchers (3,4). These principles are important for a good collaboration, but do not reflect the quality of patient engagement or its impact on outcomes of clinical research. For example, patients and researchers can have a good relationship and enjoy working together, but do not critically evaluate outcome measures, recruitment strategies, consent forms, or dissemination plans for study results.

The QUEST Center at the Berlin Institute of Health strives to increase the trustworthiness, usefulness and ethics of biomedical research (5). Engaging patients in clinical research is one approach to reach these goals. At QUEST, we intend to inform clinical research about high quality patient engagement. High quality means that the patient engagement process has the potential to increase the relevance and usefulness of clinical research for society, as well as its transparency, robustness, or ethics.

In general, quality in clinical research is assessed with critical appraisal tools, such as the widely used risk of bias tool for randomized controlled trials (RCTs) (6,7). A critical appraisal tool to assess the quality of patient engagement was developed in 2010 (10). However, in order to allow for critical appraisal and quality assessment, high quality reporting is needed. The reporting of RCTs, for instance, is still sub-optimal, but has improved since the introduction of the CONSORT reporting guideline (8–10). These evaluation studies also show that the endorsement of CONSORT by medical journals can play an important role in improving reporting quality (8–10). In the case of patient engagement, a reporting guideline (GRIPP) was developed in 2011 (11) and updated in 2017 to GRIPP2 (12). GRIPP2 comes in two different formats: a long form (LF) for studies with patient engagement as primary focus and a short form (SF) for studies with patient engagement as secondary or tertiary focus, such as for example clinical studies being informed by an active involvement of patients.

In order to analyze and compare different forms of patient engagement and their influence on the clinical study, it is necessary to report patient engagement in the publication describing the main findings of the clinical study. In 2014, the BMJ (British Medical Journal) group endorsed a policy which made it a requirement to report on patient and public involvement in BMJ journals and recommends GRIPP2 as a reporting standard (13). Price et al. (14) compared reporting of patient and public involvement before and after the introduction of the BMJ policy. They concluded that while more patient and public involvement (PPI) was reported following the introduction of the policy, only a small percentage of studies conducted patient engagement.

Funding organizations can play an important role in improving the quality and reporting of patient engagement, especially as they increasingly require patient engagement in clinical research. The NIHR, for example, refers to the GRIPP2-guideline as a resource for reporting on patient and public involvement (15). However, to our knowledge, no funding organization in the field requires the use of GRIPP2. Reporting could of course also be assessed differently. However, in order to compare and understand outcomes and methods, applying a systematically developed reporting guideline as a standard has advantages.

It is difficult to identify clinical research in which patient engagement informs study methodology, as terms vary widely and are often used with different meaning. PCORI, a US-based organization funding patient-centered research on different levels, continuously screens Medline via Pubmed, relevant journals, and PCORI staff recommendations for publications on engagement in health research (16). While many of the included publications are based on projects funded by PCORI, other publications are also included in the database.

Reviewing the literature, we found only one study, which investigated the quality of reporting or critical appraisal of patient engagement (17). Jones et al. (17) identified three studies in which patients who were not themselves participants engaged in the study. Two of them had patient engagement as primary focus, such as prioritizing research topics for colitis ulcerosa (18) or developing a patient portal (19). This is very important but differs from the scope of our study.

We will focus on studies that actively involved patients to inform all aspects of the study methodology (including dissemination) but did not have patient engagement as primary focus. Patients can be people affected by the disease or topic, or representatives of those affected. Participating in a study is not sufficient to qualify as patient engagement. We aim to analyze the extent and quality of patient engagement in best practice examples. To identify best practice examples, we will start with three samples, which we screened for the aforementioned studies (for more details see section “Samples” below):

- all publications in the category “research” published in 2019 in the British Medical Journal (BMJ)
- the PCORI health literature explorer for examples of patient engagement in 2019. Examples of patient engagement are defined by PCORI as “manuscripts with a primary objective of reporting on a health research study that engaged partners in at least one phase of the research and describe at least one impact of engagement on their work”.
- research publications citing GRIPP2 publications (12,20) and/or the critical appraisal publication by Wright et al. (21)

We aim to analyze

1. the phase of a research project, in which patient engagement was reported (short: phase of engagement)
2. the quality of reporting using GRIPP2-SF (12)
3. the quality of patient engagement using the critical appraisal tool from Wright et al. (21)

We will additionally report the frequency of patient engagement in all publications in the rubric “research” in 2019 in the BMJ, given the requirement to report whether patient and public involvement took place (13).

All included publications across these samples will be checked for additional links or references which describe patient engagement in more detail. This additional information will be checked in more depth in a sub-sample of publications. The size of this sub-sample will be adjusted depending on how many publications point to additional information. We will describe how we found further material (e.g. link provided in publication) and which additional information was found.

### Methods

#### Samples

##### *Sample 1, The BMJ 2019*

We searched Web of Science (04/03/2020) for all publications from The BMJ in 2019 with the publication type “article” or “review”. All hits were assessed independently by AS and LB. In a first step, only publications in the rubric “research” were included; research news, research methodology, etc. were excluded. In a second step, AS and LB assessed whether these studies reported patient engagement. SW assessed those with discrepant judgements and the results were discussed in the whole team. The definition of patient engagement was further refined as follows:

- Dissemination strategies to inform (participating) patients and the public without actively involving patients or patient representatives were not categorized as patient engagement.
- If dissemination strategies actively involving patients and the public were planned, but not yet conducted, this was categorized as patient engagement.
- If a patient reviewer of the BMJ has reviewed the publication, this was not categorized as patient engagement.

##### *Sample 2, PCORI 2019*

We used the “Engagement in Health Research Literature Explorer” database provided by PCORI (https://www.pcori.org/engagement/engagement-literature?f%5B0%5D=field_article_stakeholders%3A452&f%5B1%5D=field_article_topics%3A514&f%5B2%5D=field_article_date%3A2019 ). We applied the filter “example of engagement in health research” (defined as manuscripts with a *primary objective* of reporting on a health research study that engaged partners in at least one phase of the research and describe at least one impact of engagement on their work), in the category "topic", "patient" in the category "stakeholder", and "2019" in the category "year". PCORI provided us with the dataset (19/03/2020). All publications describing research were included regardless of whether they were funded by PCORI or not; protocols were excluded.

##### *Sample 3: Citations*

We conducted citation analyses for publications citing the GRIPP-2 reporting guideline (12,20) and/or the critical appraisal tool (21) in Google Scholar, Web of Science, Medline via Pubmed, and Dimensions. We chose to use Dimensions as the citation analysis resulted in the most hits and covered all three publications. After exclusion of 14 duplicates, 225 publications remained (extracted from Dimensions on 20/03/2020). We did not filter for document type at this stage. A random sample of 10 publications were assessed by the whole team before the screening process to define inclusion and exclusion criteria. LB screened the whole dataset and included publications describing research if active patient engagement informed the study methodology. Publications were excluded if none of the three original publications (12,20,21) were in fact cited, or if the publication did not describe active patient engagement in clinical research. Publications which could not be easily categorized were assessed by the whole group. Publications referring to a publication which might potentially fulfil our criteria were referred for later assessment.

#### Classification

SW, SGS, AS, LB and DF will conduct the assessment of the publications. Every publication will be assessed independently by at least two people. In case of discrepancies, a third person will solve the discrepancies by consensus, and if not possible, by majority vote.

- Phase of engagement: All included publications will be analyzed for the phase of the research project, in which patient engagement was reported: study design, study conduct and dissemination (see Table 1). In order to fully capture different aspects of study design, we added two sub-categories under "study design": "research question" and "outcome measures".
- Reporting: All included publications will be analyzed using GRIPP2-SF (see Table 2). In the pilot study, we found that it was difficult to judge unambiguously whether the criteria were fulfilled. Therefore, we decided to iteratively develop and apply additional sub-codes for the five defined criteria. To define sub-codes based on the literature, we will conduct several consensus rounds when coding the first 20 publications.
- Quality: All included publications will be assessed with the critical appraisal tool for patient engagement developed by Wright et al. (21) (see Table 3).
- Further links and references: All included publications will be searched for additional links or further references (see Table 4). In a sub-sample, we will check the information in depth and describe the additional information found and how we found it (e.g. link provided in publication).

#### Pilot Study

In a pilot study, SGS and AS analyzed five publications from The BMJ in 2019 in the rubric "research" (Sample 1), and five publications randomly selected from the PCORI health research literature explorer using the filters "examples of patient engagement", "2019", "patient" (Sample 2).

In Sample 1, two out of five publications described dissemination plans. However, neither reported that patients were involved in conceiving these dissemination plans. We decided that dissemination to participants or stakeholders will not be counted as patient engagement if patients were not "involved in choosing the methods and agreeing plans for dissemination of the study results" as described in The BMJ Patient and Public Partnership (13). According to this definition, none of the included studies conducted patient engagement.

In Sample 2, four out of five publications reported on patient engagement. One only mentioned in the acknowledgement that they thank those stakeholders involved. This was not counted as patient engagement. One of the four remaining publications was a protocol. Protocols could be very informative about the engagement of patients in the design phase. However, this protocol did not report on already conducted patient engagement, but on future activities planned with respect to patient engagement. Therefore, we decided to exclude protocols as index publications from our sample and only use them as additional sources of information for already conducted studies. The remaining three publications reported patient engagement in study design (n=3), study conduct (n=2), and dissemination (n=1). Two publications reported patient engagement in defining the research question, and three in determining outcome measures. Two publications gave some information with respect to the aim of patient engagement in the study, three with respect to the methods used for patient engagement. One publication gave some information about positive and negative outcomes of patient engagement, another about the extent to which patient engagement influenced the study overall, and two reflected somewhat critically on the patient engagement. However, coding for all categories was rather vague and dependent on the interpretation of sentences or half-sentences. Based on this result of the pilot studies, we decided to iteratively develop sub-codes for the five reporting categories of GRIPP2-SF to describe and define them in more detail.

There was very limited reporting on the impact of patient engagement on the study methodology. This made it difficult to critically appraise the publication using the checklist by Wright et al. In two of the three publications we identified some information about the rationale for patient engagement. In the third publication, the authors mentioned how they recruited the patient for one part of the patient engagement. Unfortunately, there was no information in any of the publications about the appropriateness of the involvement, training, ethical and/or methodological interventions, dissemination, evaluation and impact assessment. Nevertheless, we decided to assess all publications with the checklist.

Further links or references were given in three of the five publications. Additional information was available for all of them on the PCORI database as they were all funded by PCORI.

#### Codebook

*Table 1: Phase of Engagement*

| Code | Description | Rationale |
|---|---|---|
| **Study Design** | Including<br>- design of intervention<br>- advisory board<br>- ..<br>Excluding<br>- research question<br>- outcome measure | Patient engagement in study design and planning should have the greatest impact on conducting more patient relevant research, especially for rigorous study designs, such as RCT. |
| **Research Question** | Only if specifically mentioned, otherwise code as “study design” | Patient engagement in defining, prioritizing, and describing the research question might have the biggest impact on outcomes in clinical research. |
| **Outcome measures** | Only if specifically mentioned, otherwise code as “study design” | Patient engagement in prioritizing outcome measures is highly important for increasing the value of the research. |
| **Study Conduct** | including<br>- recruitment | Depending on the study methodology, the impact of patient engagement during |

| Code | Description | Rationale |
|---|---|---|
| | - advisory board during study conduct<br>- analysis<br>- interpretation of results<br>- drafting of the manuscript<br>- .. | study conduct can vary widely. There will be much less possibilities for impact in more rigorous study designs, such as RCTs. |
| **Dissemination** | Only if patients were involved in choosing methods and agreeing plans for dissemination | Patient engagement in dissemination of outcomes of clinical research might not influence research itself, but its success in translating into health care. |

*Table 2: Reporting (GRIPP2-SF, see Staniszewska et al. (12,20)).*

| Code | Definition |
|---|---|
| **Aim** | Report the aim of PPI in the study |
| **Methods** | Provide a clear description of the methods used for PPI in the study |
| **Study Results** | Outcomes—Report the results of PPI in the study, including both positive and negative outcomes |
| **Discussion and conclusions** | Outcomes—Comment on the extent to which PPI influenced the study overall. Describe positive and negative effects. |
| **Reflections/critical perspective** | Comment critically on the study, reflecting on the things that went well and those that did not, so others can learn from this experience |

As the reporting items are not very specifically defined and were difficult to code in the pilot study (see above), we decided to create sub-codes in order to further specify and define the codes.

*Table 3: Critical Appraisal (see Wright et al. (21))*

| Topic | Question | Consider the following |
|---|---|---|
| **Planning and project design** | 1. Is the rationale for involving users clearly demonstrated? | (a) Have the researchers explained the rationale for user involvement? |
| | 2. Is the level of user involvement appropriate? | (a) Have the researchers explained and justified the level of user involvement |
| | | (b) Have the researchers discussed the nature of tasks users were asked to perform (e.g. identifying the research question, selecting the research method, commenting on information sheets, data collection, data analysis, dissemination?) |
| **Recruitment and training** | 3. Is the recruitment strategy appropriate? | (a) Have the researchers explained how users have been identified? |
| | | (b) Have attempts been made to involve a wide cross-section of interests where appropriate (e.g. ethnic minorities, age, gender)? |
| | | (c) Have the researchers discussed the credentials of the users involved? (E.g. Do the researchers discuss why the users involved are appropriate to meeting the aims of the involvement activity?) |
| | 4. Is the nature of training appropriate? | (a) Have the researchers discussed the nature of the training provided? |
| | | (b) Is the nature and extent of the training justified by the researchers? (e.g. Do the researchers |

| Topic | Question | Consider the following |
|---|---|---|
| | | discuss how the training meets the needs of the users during the course of the study?) |
| | | (c) Has an account been given of user involvement training for professional researchers, where necessary? |
| **Data collection and analysis** | 5. Has sufficient attention been given to the ethical considerations of user involvement and how these were managed? | (a) Do the researchers discuss ethical issues relating to the involvement of users in research (e.g. fatigue, the emotional demands of data collection)? |
| | | (b) Are there any discussions about the management of ethical issues (e.g. provision of adequate information about research tasks, peer supervision)? |
| **Data collection and analysis** | 6. Has sufficient attention been given to the methodological considerations of user involvement and how these were managed? | (a) Have the researchers discussed methodological issues relating to user involvement in research (e.g. potential impact on the quality of the data)? |
| | | (b) Do the researchers discuss how methodological issues are managed (e.g. how differences in interpretations of qualitative data are negotiated?) |
| **Dissemination** | 7. Have there been any attempts to involve users in the dissemination of findings? | (a) Have users been involved in the writing of the publication / funding application? |
| | | (b) Have the researchers described how the findings have been disseminated to participants and service users? |
| | | (c) Are findings disseminated appropriately where necessary (e.g. translation of findings into different languages, provision of interim findings to participants in receipt of palliative care)? |
| **Evaluation and impact assessment** | 8. Has the added-value of user involvement been clearly demonstrated? | (a) Do the researchers discuss what difference involving users in the design and conduct of the research has made to the research process? (I.e. Have the researchers considered whether the study and findings would look any different if users were not involved?) |
| | | (b) Do the researchers support the claims for the benefits of user involvement with examples from the research project? |
| | 9. Have there been any attempts to evaluate the user involvement component of the research? | (a) Have the researchers discussed the evaluation of the impact of user involvement on the research project (e.g. impact on the length of the study, the financial cost of involvement activities, cost-benefit analyses)? |

| Topic | Question | Consider the following |
|---|---|---|
| | | (b) Do the researchers support claims about the impact of user involvement with examples from the evaluation? |

*Table 4: Further Links or References*

| Code | Definition |
|---|---|
| **Further links or references available** | - Are there links or references mentioned in the text in which the patient engagement is described in more detail?<br>- Are there more links or references provided in the PCORI database? |

For a sub-sample of publications, we will check the additional links and references in more depth. We will describe how we found further material (e.g. link provided in publication) and which additional information we found.

#### Author contributions

SGS, AS, SW, DF and DS designed the study. SGS and AS conducted the pilot study. LB and AS independently screened the BMJ sample, disagreements were discussed and consented between all authors. LB screened the citation sample. SGS wrote the first draft of the protocol, all authors revised and approved it.

#### Funding

The study is funded by intramural funds of the QUEST Center, Berlin Institute of Health, Charité – Universitätsmedizin Berlin.

### Supplement 2: Targeted Search and Flowchart

#### Search Strategy

##### BMJ Sample

- Database: Web of Science Core Collection
- Extracted on March 4, 2020
- Search query: IS=1756-1833 AND PY=2019 (document types: 'Article' or 'Review').
- Hits: 200 publications

##### PCORI Sample

- Database: PCORI health literature explorer (https://www.pcori.org/engagement/engagement-health-research-literature-explorer/engagement-health-research-literature-explorer-supplemental-methods-information)
- Extracted on March 19, 2020
- Filter:
  - topic: example of engagement in health research*;
  - stakeholder involvement: patients;
  - year: 2019.
- Hits: 98 publications

*"Examples of Engagement in Health Research" are defined by PCORI as "manuscripts with a primary objective of reporting on a health research study that engaged partners in at least one phase of the research and describe at least one impact of engagement on their work.

##### Citation Sample

- Database: Dimensions*
- Extracted on March 20, 2020
- Citation analyses for Staniszewska et al. (2017a, 2017b) and Wright et al. (2010)
- Hits: 239 publications (after exclusion of 14 duplicates, 225 publications remained).
  - 195 papers cited the GRIPP2 publications
  - 44 cited the critical appraisal publication

*we conducted analysis in Web of Science, Scite, Medline via PubMed, Google Scholar, and Dimensions. We chose to use Dimensions as the citation analysis resulted in the most hits and covered all three reference publications.

#### Flowchart: Publication screening

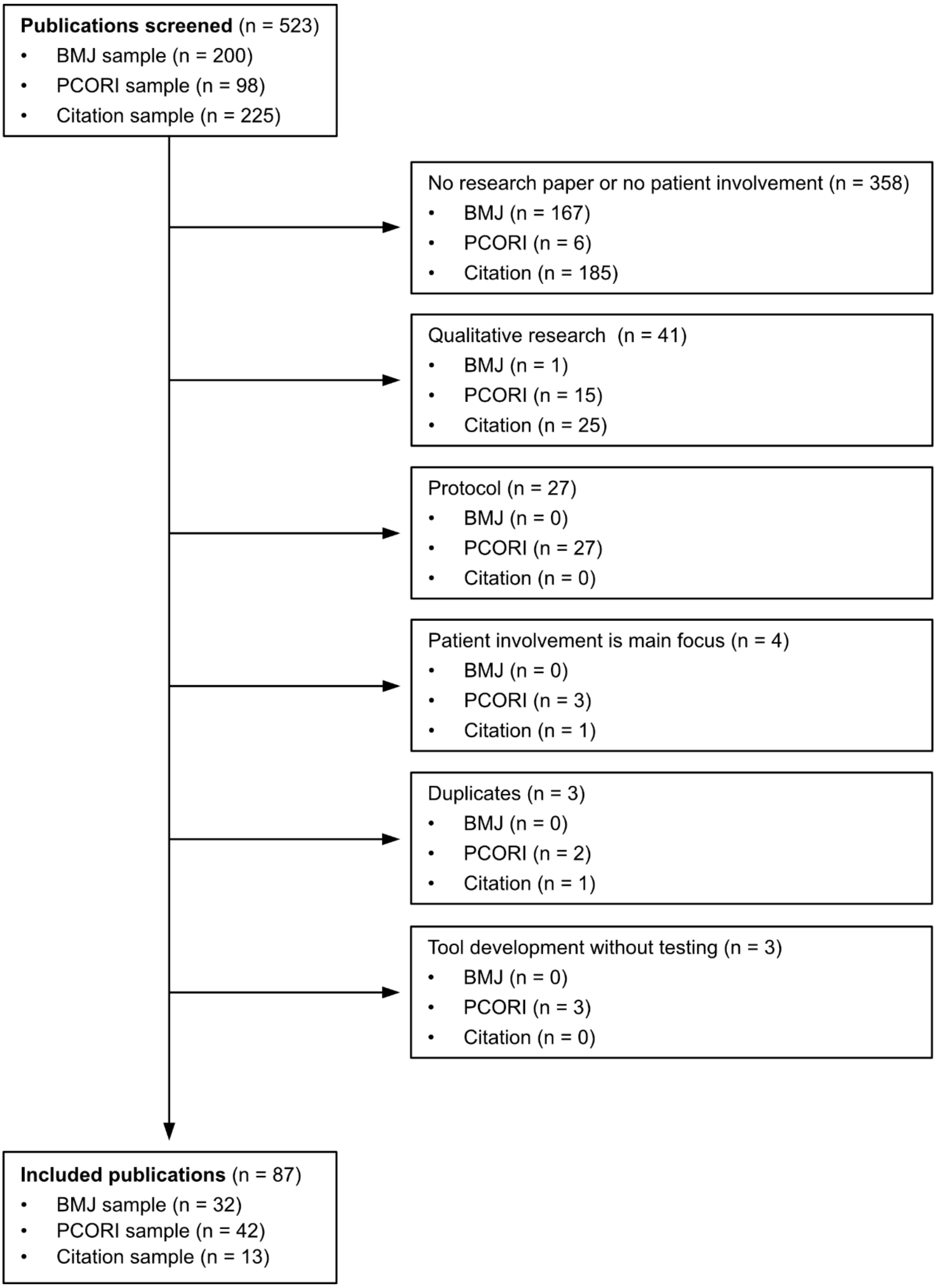

#### Supplement 3: Coding Schemes

*Table S3.1: Phase of involvement.*

| Code | Description | Rationale |
|---|---|---|
| **Study design** | Including<br>- design of intervention<br>- advisory board | Patient involvement in study design and planning should have the greatest impact on conducting more patient relevant research, especially for rigorous study designs, such as RCT. |
| • **Research question**[1] | Only if specifically mentioned, otherwise code only as "study design" | Patient involvement in defining, prioritizing, and describing the research question might have the biggest impact on outcomes in clinical research. |
| • **Outcome measures**[1] | Only if specifically mentioned, otherwise code only as "study design" | Patient involvement in prioritizing outcome measures is highly important for increasing the value of the research. |
| **Study conduct** | Including<br>- recruitment<br>- advisory board during study conduct<br>- analysis<br>- interpretation of results | Depending on the study methodology, the impact of patient involvement during study conduct can vary widely. There will be much less possibilities for impact in more rigorous study designs, such as RCTs. |
| **Dissemination** | Only if patients were involved in choosing methods and agreeing plans for dissemination | Patient involvement in dissemination of outcomes of clinical research might not influence research itself, but its success in translating into health care. |
| • **Co-Authoring the manuscript**[1,2] | If patient partner was one of the authors | |

1 Deviating from the protocol, these subcodes were coded in addition to the main code.
2 This subcode was not specified in the protocol, but added during the process.

*Table S3.2: GRIPP2 reporting guidelines short form according to Staniszewska et al. (Staniszewska et al., 2017a, 2017b).*

| Code | Definition | Further specification (developed during coding) |
|---|---|---|
| **Aim** | Report the aim of PPI in the study | |
| **Methods** | Provide a clear description of the methods used for PPI in the study | |
| **Study Results** | Outcomes—Report the results of PPI in the study, including both positive and negative outcomes | concrete and detailed outcomes of patient involvement (e.g., specific comments of patients, which led to a change of an intervention) |
| **Discussion and conclusions** | Outcomes—Comment on the extent to which PPI influenced the study overall. Describe positive and negative effects. | general outcomes of patient involvement (e.g., statements about enhancing the validity of the study by patient involvement) |
| **Reflections/critical perspective** | Comment critically on the study, reflecting on the things that went well and those that did not, so others can learn from this experience | |

*Table S3.3: Critical appraisal tool according to Wright et al. (2010).*

| Question | Consider the following | Further specification (developed during coding) |
|---|---|---|
| **Planning and project design** | | |
| 1. Is the rationale for involving users clearly demonstrated? | (a) Have the researchers explained the rationale for user involvement? [Rationale] | |
| 2. Is the level of user involvement appropriate? | (a) Have the researchers explained and justified the level of user involvement [Level of involvement] | |
| | (b) Have the researchers discussed the nature of tasks users were asked to perform (e.g. identifying the research question, selecting the research method, commenting on information sheets, data collection, data analysis, dissemination?) [Nature of tasks] | |
| **Recruitment and training** | | |
| 3. Is the recruitment strategy appropriate? | (a) Have the researchers explained how users have been identified? [Identification] | |
| | (b) Have attempts been made to involve a wide cross-section of interests where appropriate (e.g. ethnic minorities, age, gender)? [Diversity] | |
| | (c) Have the researchers discussed the credentials of the users involved? (E.g. Do the researchers discuss why the users involved are appropriate to meeting the aims of the involvement activity?) [Credentials] | |
| 4. Is the nature of training appropriate? | (a) Have the researchers discussed the nature of the training provided? [Nature of training] | |
| | (b) Is the nature and extent of the training justified by the researchers? (e.g. Do the researchers discuss how the training meets the needs of the users during the course of the study?) [Justification of the training] | |
| | (c) Has an account been given of user involvement training for professional researchers, where necessary? [User involvement training for researchers] | |
| **Data collection and analysis** | | |
| 5. Has sufficient attention been given to the ethical considerations of user involvement and how these were managed? | (a) Do the researchers discuss ethical issues relating to the involvement of users in research (e.g. fatigue, the emotional demands of data collection)? [Ethical issues] | |
| | (b) Are there any discussions about the management of ethical issues (e.g. provision of adequate information about research tasks, peer supervision)? [Management of ethical issues] | |
| 6. Has sufficient attention been given to the methodological considerations of user involvement and how these were managed? | (a) Have the researchers discussed methodological issues relating to user involvement in research (e.g. potential impact on the quality of the data)? [Methodological issues] | |
| | (b) Do the researchers discuss how methodological issues are managed (e.g. how differences in interpretations of qualitative data are negotiated?) [Management of methodological issues] | |
| **Dissemination** | | |
| 7. Have there been any attempts to involve users in the dissemination of findings? | (a) Have users been involved in the writing of the …funding application? **[**Writing of the funding application] | Code was split into two subcodes |
| | .. of the publication? [Writing of the publication] | |

| | | |
|---|---|---|
| | (b) Have the researchers described how the findings have been disseminated to participants and service users? [Description of dissemination] | |
| | (c) Are findings disseminated appropriately where necessary (e.g. translation of findings into different languages, provision of interim findings to participants in receipt of palliative care)?[ Accessible dissemination] | |
| **Evaluation and impact assessment** | | |
| 8. Has the 'added-value' of user involvement been clearly demonstrated? | (a) Do the researchers discuss what difference involving users in the design and conduct of the research has made to the research process? (I.e. Have the researchers considered whether the study and findings would look any different if users were not involved?) [Difference made to the research process] | |
| | (b) Do the researchers support the claims for the benefits of user involvement with examples from the research project? [Examples of assessment of the benefits] | |
| 9. Have there been any attempts to evaluate the user involvement component of the research? | (a) Have the researchers discussed the evaluation of the impact of user involvement on the research project (e.g. impact on the length of the study, the financial cost of involvement activities, cost-benefit analyses)? [Evaluation of the made impact] | Only those were coded which conducted a more formal evaluation |
| | (b) Do the researchers support claims about the impact of user involvement with examples from the evaluation? [Examples of evaluation] | |
